# A Hybrid Rule-Based and Deep Learning Framework for Ventilator Waveform Segmentation and Delineation

**DOI:** 10.64898/2026.01.24.26344749

**Authors:** Sagar Deep Deb, Suvin Shetty, Arpita Dwivedy, Deepak K. Agrawal

**Affiliations:** Department of Biosciences and Bioengineering, Indian Institute of Technology Bombay, Mumbai, Maharashtra, India; Dr. L. H. Hiranandani Hospital, Mumbai, Maharashtra, India

## Abstract

Accurate assessment of patient-ventilator interaction is critical for optimizing respiratory support and detecting harmful dyssynchronies linked to adverse outcomes, including ventilator-induced lung injury and prolonged ICU stays. This requires precise, breath-by-breath segmentation and phase delineation of ventilator waveforms, specifically pressure, flow, and volume. Current reliance on manual annotation limits scalability and consistency, particularly given the variability of waveforms across diverse patient conditions and ventilator settings. To address this challenge, we present a fully automated, two-stage hybrid pipeline that integrates a rule-based algorithm with a Deep Learning (DL) model. The rule-based module generates pseudo-labels by detecting steep rises in the pressure derivative for breath segmentation and analyzing zero-crossings in the flow signal for phase delineation. These labels train a modified 1D U-Net enhanced with Bidirectional Long Short-Term Memory (Bi-LSTM), which captures temporal dependencies and improves adaptability to complex waveform morphologies, such as double-triggered ventilator dyssynchrony breaths. The framework was developed using data from adult ICU patients and evaluated on an independently annotated test set. The Bi-LSTM U-Net model achieved a Dice score of 0.9611, surpassing both the rule-based method, which scored 0.9321, and baseline U-Net architectures, which scored 0.9587. The model demonstrated high temporal precision, with inspiration offset and onset errors of 0.004± 0.013 seconds and 0.013 ±0.028 seconds, respectively. The Bi-LSTM architecture proved particularly effective, reducing inspiration offset errors by 43% and onset errors by 28% compared to the rule-based method and baseline U-Net, while also maintaining low error variability. This hybrid approach provides a scalable, accurate, and fully automated solution for ventilator waveform analysis, enabling enhanced assessment of patient-ventilator synchrony without manual intervention.

## 2 Introduction

During invasive mechanical ventilation (IMV), the ventilator generates pressure, flow, and volume waveforms that are used to assess the respiratory condition and evaluate the effectiveness of interventions [1, 2]. These waveforms provide valuable insights into lung mechanics, patient–ventilator interactions, and the overall state of the respiratory system [3, 4]. As such, they serve as a foundation for tailoring ventilator settings to individual patient needs and, in doing so, minimizing complications such as ventilator-induced lung injury (VILI), optimizing gas exchange, and improving clinical outcomes [5, 6]. One key application is the detection of ventilator dyssynchrony (VD), defined as a mismatch between the spontaneous efforts of the patient and ventilator support [7]. VD is linked to adverse outcomes, including a higher risk of VILI and increased mortality [8, 9]. Identifying VD requires monitoring waveform abnormalities relative to the morphology of the normal waveform. This has led to the development of various methods, from rule-based systems [**?**, 10–13] to deep learning models [15–20], to automate offline VD detection. However, many existing approaches still rely on manual breath-by-breath segmentation, limiting scalability and reliability.

Developing a scalable automated VD detection pipeline requires precise identification of breath boundaries, such as the onset and termination of inspiration and expiration [13]. Such precise breath-level marking serves as the basis for analyzing temporal and morphological features associated with dyssyn-chrony events [13,21,22]. Therefore, accurate boundary detection could enable consistent identification of dyssynchrony patterns such as double triggering, ineffective efforts, or premature cycling, and might even support quantification of dyssynchrony severity [7,10]. Breath segmentation identifies the start of each respiratory cycle, while phase delineation distinguishes between inspiratory and expiratory phases from continuous waveform signals [23–25]. However, achieving this precision algorithmically remains a significant challenge.

Early attempts to automate breath segmentation employed rule-based algorithms applied to flow signals across ventilator modes [13]. While innovative, this approach fails to accommodate variability in VD waveform signals, such as double triggering, where back-to-back inspiratory efforts can cause brief flow reversals, leading to misidentification of the end-of-inspiration phase [4, 26]. More recently, deep learning (DL) models have been applied to improve the accuracy of breath segmentation. For example, Bakkes et al. [19] proposed a U-Net–based model, framing the task of phase delineation as a three-class sequence-labeling problem using esophageal pressure (*P*_es_) as ground truth. Although *P*_es_ offers a direct and reliable measure of patient effort, it requires invasive catheter placement and specialized handling, making it impractical for routine bedside use [27–29].

Subsequent research has investigated non-invasive methods using standard ventilator waveforms, including airway pressure (referred to as “pressure”) and airflow (referred to as “flow”). Mojoli et al. [25] showed that it is possible to detect the beginning and end of the inspiratory phase with these pressure and flow waveforms. However, their approach depended on fixed thresholds and heuristic rules, which can be affected by noise, variations in patient-ventilator interactions, and waveform artifacts, making it less effective across different waveform patterns. Despite these advancements, there remains a significant need for adaptive, patient-specific approaches that can accurately segment breaths across diverse waveform patterns [30]. Such an approach should be generalizable to accommodate the high variability observed in ventilator waveforms and scalable enough to handle large amounts of data without requiring manual annotation.

Rule-based methods employ fixed thresholds and logical rules derived from domain knowledge. They are simple to interpret and perform well on clean signals, but their accuracy drops with noisy or complex waveforms [31]. DL models can learn detailed shape and temporal features, handling more variability, but they require large annotated datasets and offer less interpretability [32]. A promising solution may lie in combining both approaches: a rule-based module provides physiologically meaningful labels, while a DL model learns to handle cases where rules fail. In this paper, we present an automated pipeline for breath segmentation and delineation that uses continuous pressure and flow waveforms to precisely identify individual breaths and their phases. This approach combines rule-based algorithms based on pressure and flow derivatives with a modified U-Net model enhanced with Bidirectional Long Short-Term Memory (Bi-LSTM) layers to effectively capture temporal dependencies. This integrated approach aims to improve both accuracy and adaptability across diverse respiratory waveforms.

The paper is organized as follows: Section 3 provides a comprehensive description of the proposed methodology. It begins with details on the study population and data collection in Subsection 3.1, followed by a formal definition of the segmentation and delineation problem in Subsection 3.2. The mathematical construction of the rule-based algorithms for breath segmentation and phase delineation is presented in Subsection 3.3. The DL model architecture is formulated in Subsection 3.4, and the processes for data selection and ground truth annotation are detailed in Subsections 3.5 and 3.6, respectively. Section 4 presents the results of both the quantitative and qualitative evaluations of the proposed framework. Section 5 examines the findings and their implications, while also outlining the limitations and future directions for research. Finally, Section 6 summarizes the key contributions and conclusions of this research.

## 3 Methods

### 3.1 Study Population and Data Collection

This study enrolled adult Intensive Care Units (ICU) patients at Dr. L.H. Hiranandani Hospital (LHHH), Mumbai, who required invasive mechanical ventilation and either met Berlin ARDS criteria or were at risk for ARDS [33,34]. Participants were aged 18 to 89 years, and enrollment occurred within 24 hours of intubation. “At-risk” was defined as a clear inciting lung injury without full radiographic or oxygenation criteria. Exclusions included age below 18, pregnancy, imprisonment, contraindications to esophageal monitoring, facial fractures, or recent bleeding diathesis. All patients were ventilated on a Maquet Servo-i. Institutional ethics approvals were obtained from LHHH and the Indian Institute of Technology Bombay.

Baseline data were collected at the start, including age, gender, height, and P/F ratio. A laptop interfaced with the ventilator acquired airway pressure, flow, and volume waveforms using Vital Recorder, along with timestamps and periodic ventilator parameters, including positive end-expiratory pressure (PEEP), tidal volume (TV), respiratory rate (RR), and oxygen concentration. Data collection continued until extubation. This analysis focused on two patients in whom the primary mode of ventilation was Synchronized Intermittent Mandatory Ventilation (SIMV). Patient PI0001 (female, 72 years) had 20 hours of recorded data from which a representative subset of 7.8 hours (7,000 breaths) was used to generate rule-based pseudo-labels for training. Patient PI0002 (male, 71 years) had 16 hours of recorded data from which a 0.57-hour subset (533 breaths) was manually annotated for independent testing.

### 3.2 Physiological Basis for Breath Segmentation and Delineation

Normal waveforms typically align patient effort with ventilator support, but common ventilatory disturbances, such as early cycling and delayed triggering, disrupt this pattern [4, 35]. These waveform changes enable accurate estimations of inspiratory onset and termination without esophageal pressure. Following the methodology of Mojoli et al. [25], our pipeline employs airway pressure and flow signals for breath segmentation and delineation (see Fig. 1(a–c)). The onset of inspiratory effort is indicated by a negative deflection in airway pressure and a positive deflection in flow (*p*_1_, *f*_1_), quickly followed by a steep rise in both signals as ventilator support begins. The end of inspiration is marked by flow decay and a downward shift in airway pressure (*p*_2_, *f*_2_), signifying inspiratory muscle relaxation [25].

**Figure 1.**
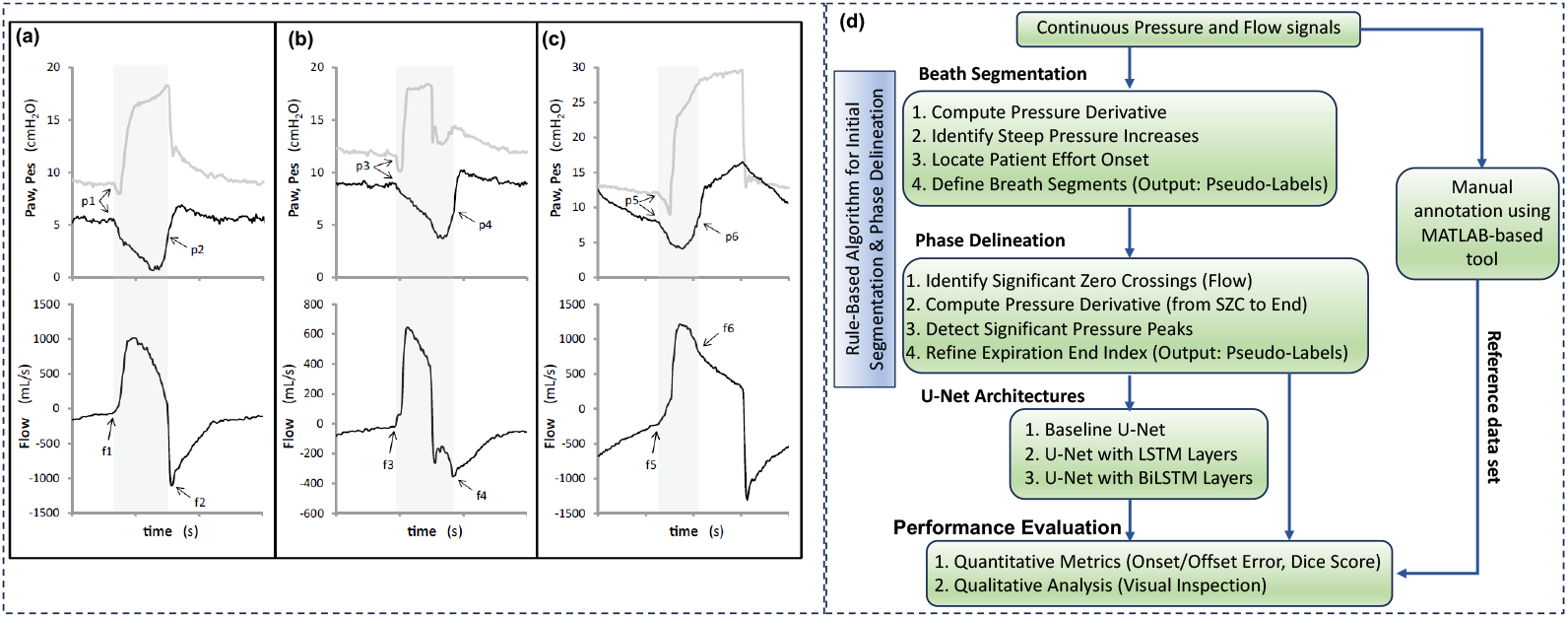
Overview of the hybrid framework for ventilator waveform segmentation and delineation. Panels (a), (b), and (c) depict different respiratory patterns: normal breathing, early cycling, and delayed trigger, respectively. At the top, airway pressure (P_aw_, in gray) and esophageal pressure (P_es_, in black) are presented, while flow is shown below. The gray-shaded areas indicate the neural inspiratory time derived from P_es_. Inspiration begins with negative deflections in both P_aw_ and P_es_ (*p*_1_, *p*_3_, *p*_5_), accompanied by positive deflections in flow (*f*_1_, *f*_3_, *f*_5_). The conclusion of the inspiratory phase typically aligns with the midpoint of the rapid rise in P_es_ (*p*_2_, *p*_4_, *p*_6_) and the initial phase of exponential decay in flow (*f*_2_, *f*_4_, *f*_6_). Reproduced from Mojoli et al., licensed under CC BY 4.0 [25]. Panel (d) outlines the proposed pipeline that combines a rule-based method with a deep learning (DL) model for breath segmentation and phase delineation.

In optimal conditions, the peak expiratory flow is immediately followed by an exponential decay (Fig. 1(a)). When cycling occurs early, the exponential decay of expiratory flow is delayed, marking the end of inspiration during expiration (Fig. 1(b)). Conversely, delayed cycling results in exponential decay occurring during inspiration after a change in slope (Fig. 1(c)).

Our pipeline leverages identifiable features to implement a two-stage process (Fig. 1(d)). In the first stage, a rule-based algorithm segments and delineates continuous pressure and flow signals by (i) identifying individual breaths through amplified rapid positive deflections in the differentiated pressure waveform and (ii) distinguishing inspiration and expiration via zero-crossings and sign changes in flow signals. This process generates pseudo-labels. In the second stage, these labels are used to train various DL models, including a baseline U-Net, a U-Net with LSTM, and a U-Net with Bi-LSTM, followed by performance evaluation.

### 3.3 Mathematical construction of Rule-Based Framework

We apply the physiological insights detailed in the previous subsection through a rule-based algorithm designed to identify crucial waveform events systematically. The algorithms for breath segmentation and delineation are illustrated in Fig. 2 (a) and (b), respectively.

**Figure 2.**
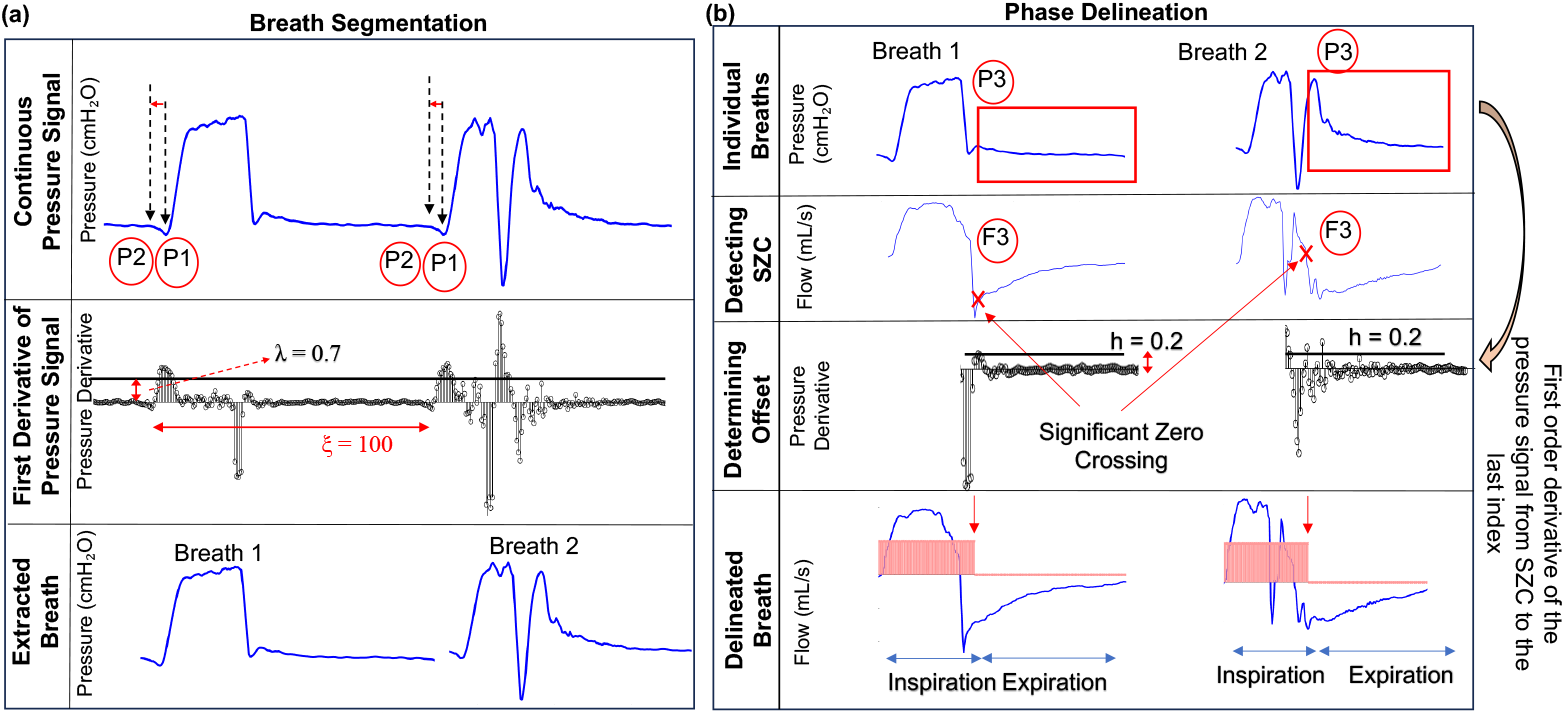
Rule-based algorithm for breath segmentation and phase delineation. Panel (a) illustrates breath segmentation, showing pressure waveforms with key markers: P1 for the initiation of ventilator support and P2 for the onset of patient effort, followed by the first derivative of the pressure signal with a threshold (*λ*) and an inter-breath interval parameter (*ξ*), along with examples of segmented breaths. Panel (b) details phase delineation using segmented pressure and flow waveforms, with F3 estimating the end of inspiration and P3 (the pressure derivative) refined by a threshold (*h*), along with examples of breaths with delineated inspiratory phase (pink-shaded region).

The method begins by detecting the notable pressure increase that indicates the initiation of ventilator support (P1) by identifying peaks in the pressure derivative that surpass an adaptive threshold, *λ* (Fig. 2a). Following each P1 detection, the algorithm conducts a backward search to locate the preceding negative deflection that signifies the onset of patient effort (P2). The threshold *λ* is customizable for each patient, accounting for variations in lung compliance and ventilator settings. To maintain physiological validity, the algorithm mandates a minimum window interval, *ξ* (number of data points), between consecutive P2 points, thereby preventing false detections caused by signal noise or double-trigger events. Here, the F1 and F2 points in the flow signal relative to the pressure signal were omitted, as they were not used during breath segmentation.

The phase delineation process, as shown in Fig. 2(b), assesses the onset of expiration through flow zero-crossing analysis. The algorithm initially identifies significant zero-crossings (SZCs) in the segmented breath flow signal, denoted F3, and in the pressure signal, denoted P3. We filter these to keep only “significant” zero crossings, those with no other crossings within a window of *w* (number of data points), reducing the chance of false positives from noise. In cases of VD, flow may turn negative before inspiration actually ends. To differentiate genuine expiratory onset from transient fluctuations, we evaluate the pressure derivative immediately after each candidate zero-crossing. A critical insight is that values exceeding the threshold *h* signify confirming pressure spikes that reliably indicate the onset of expiration. If no such spike is found, the algorithm conservatively designates the zero-crossing point F3 as the expiratory transition. This dual-signal verification can improve the accuracy of phase transition identification despite waveform variability.

#### 3.3.1 Breath segmentation based on steep rise in pressure signal

Breath onsets are identified by detecting steep rises in the pressure signal *p*[*n*] that exceed the threshold *λ*. For that, we compute the discrete derivative of the pressure signal, represented as *y*[*n*]:

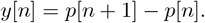

Here, *n* denotes discrete time indices. We establish a set ℐ of candidate breath-start indices where the derivative exceeds the threshold:

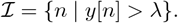

For each of the *i*^*th*^ candidate in set ℐ, we look backward to find an index *d* marking a sudden negative deflection:

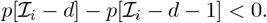

The refined effort onset candidate is:

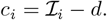

The set *C* = *c*_1_, *c*_2_, …, *c*_*k*_ includes all such points. To avoid false positives, we enforce a minimum interval *ξ* between detections, resulting in the set:

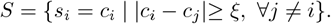

The start of breath (*k* + 1)^th^ implicitly marks the end of breath *k*^th^, eliminating explicit endpoint detection. The algorithm outputs the start and end indices of the pressure signal, defining each breath segment and its corresponding flow signal. Initial pressure data (indices 1–20) are excluded; analysis begins at index 21. The step-by-step procedure is detailed in Algorithm 1.

##### Algorithm 1

Breath segmentation based on steep rise in pressure signal

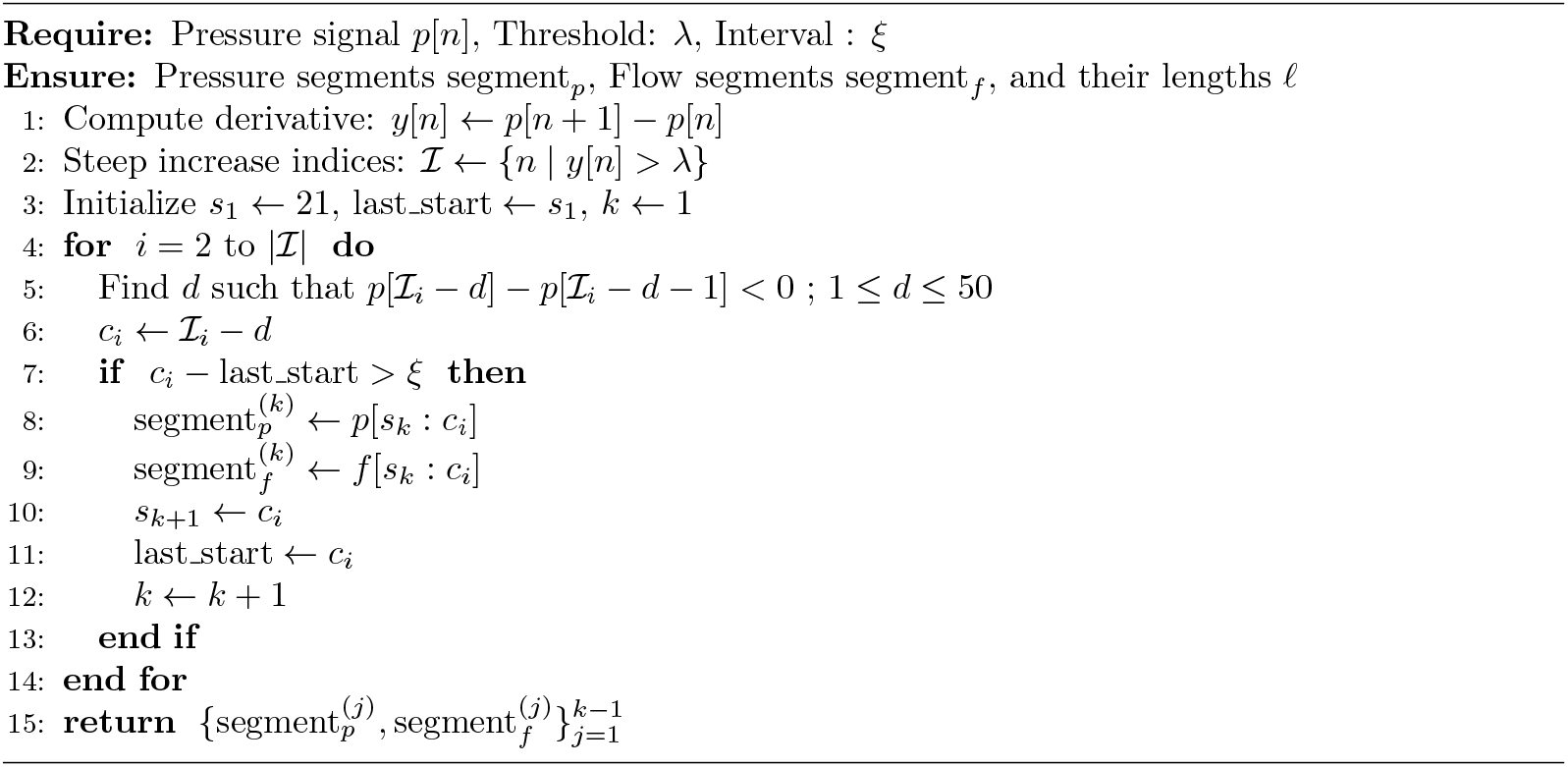

#### 3.3.2 Breath phase delineation based on zero crossing of flow signal

To determine the end of inspiration, we analyze flow and pressure signals within each breath segment. Let *f* [*n*] represent the flow signal in each segment. We identify SZCs *T* in *f* [*n*] where the derivative changes sign, and filter them to reduce noise by requiring a minimum separation *w* between crossings.

The last significant zero-crossing *e* in *T* serves as the tentative expiratory onset. For cases of VD where flow may turn negative prematurely, we analyze the pressure derivative Δ*y* after *e*:

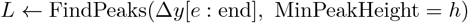

The first peak in *L* indicates residual pressure activity, and its position refines the inspiration offset. This combined approach of flow zero-crossing and pressure-peak detection improves breath-phase delineation. The detailed step-by-step methodology is outlined in Algorithm 2, which returns *l*(*n*), the delineation labels corresponding to *f* [*n*].

##### Algorithm 2

Breath Phase Delineation Using Flow and Pressure Signals

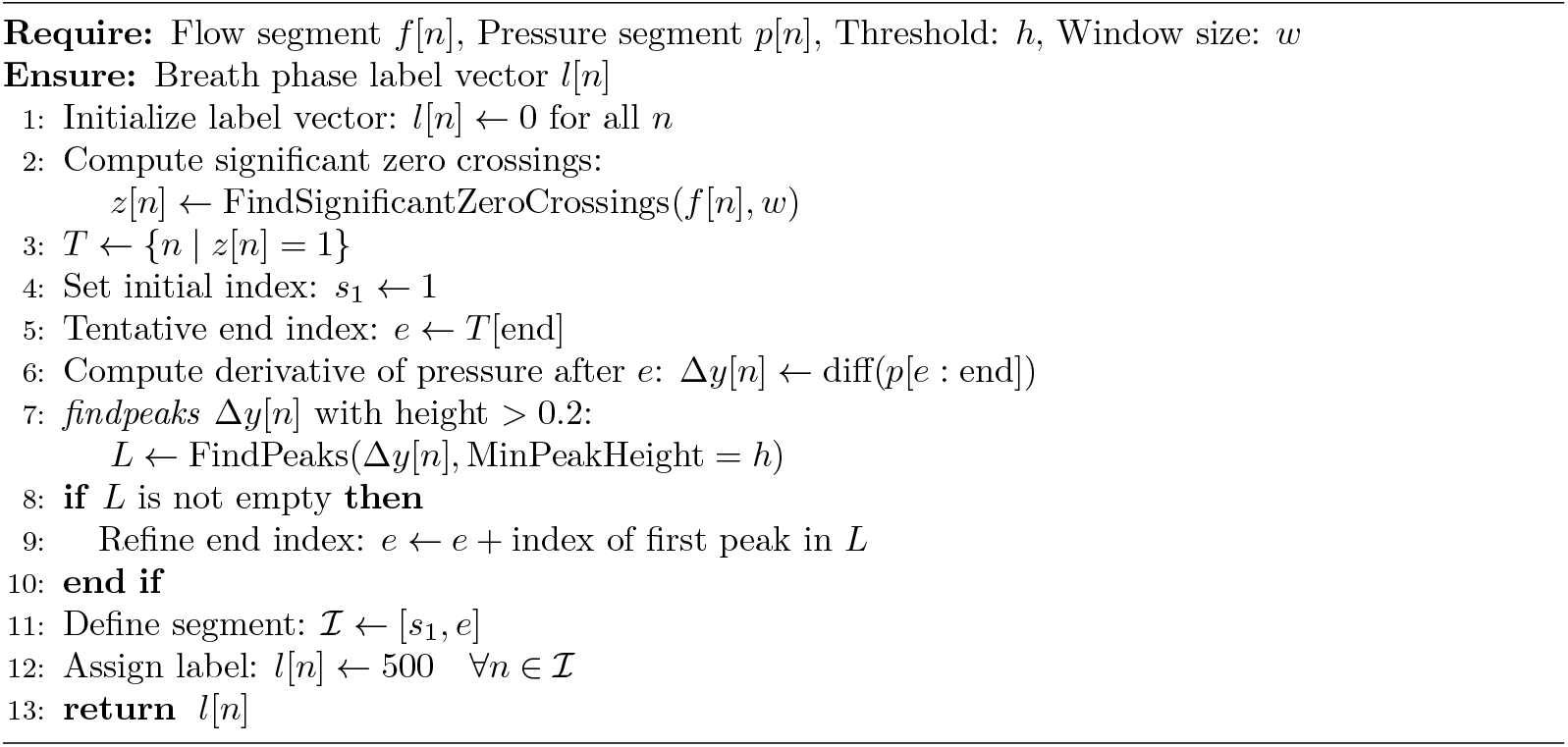

### 3.4 Deep Learning Model Formulation

To address the limitations of rule-based methods, which are heavily influenced by waveform variability, we adopt a hybrid strategy (Fig. 1(d)). This approach uses rule-based outputs as automated preliminary labels to train a deep learning model, facilitating more adaptive and precise breath segmentation and phase delineation compared to traditional static threshold techniques. The model processes concatenated flow signals, which offer the most direct and immediate measure of pulmonary air movement, yielding clear inspiratory and expiratory phase transitions [13].

We investigate various architectural configurations within the bottleneck of our 1D U-Net model to enhance the capture of long-range temporal dependencies (see Fig. 3). The architecture is adapted from the traditional U-Net framework [36], featuring an encoder-decoder structure with skip connections. These connections are critical as they enable the model to retain both global context and fine-grained details, making the design particularly effective for processing one-dimensional signals such as ECG and lung sounds [37,38]. To further optimize the U-Net for ventilator waveform segmentation, we integrate Long Short-Term Memory (LSTM) and Bidirectional LSTM (Bi-LSTM) layers into the decoder. This enhancement significantly improves the model’s capability to capture temporal dependencies and phase transitions inherent in respiratory waveforms [39, 40]. The network is organized into three encoder-decoder blocks, each utilizing (3 × 1) convolutional kernels. In the encoder, max pooling is applied for effective downsampling, while the decoder employs 2× upsampling layers, facilitating multi-resolution feature extraction. Skip connections play a vital role in preserving spatial and contextual information as it flows through the network [41, 42]. Moreover, LSTM and Bi-LSTM layers, each consisting of 64 units, are incorporated into the architecture to adeptly capture complex sequential patterns in the respiratory data [43, 44]. Ultimately, the model processes the input flow signal and produces a delineated breathing pattern as output.

**Figure 3.**
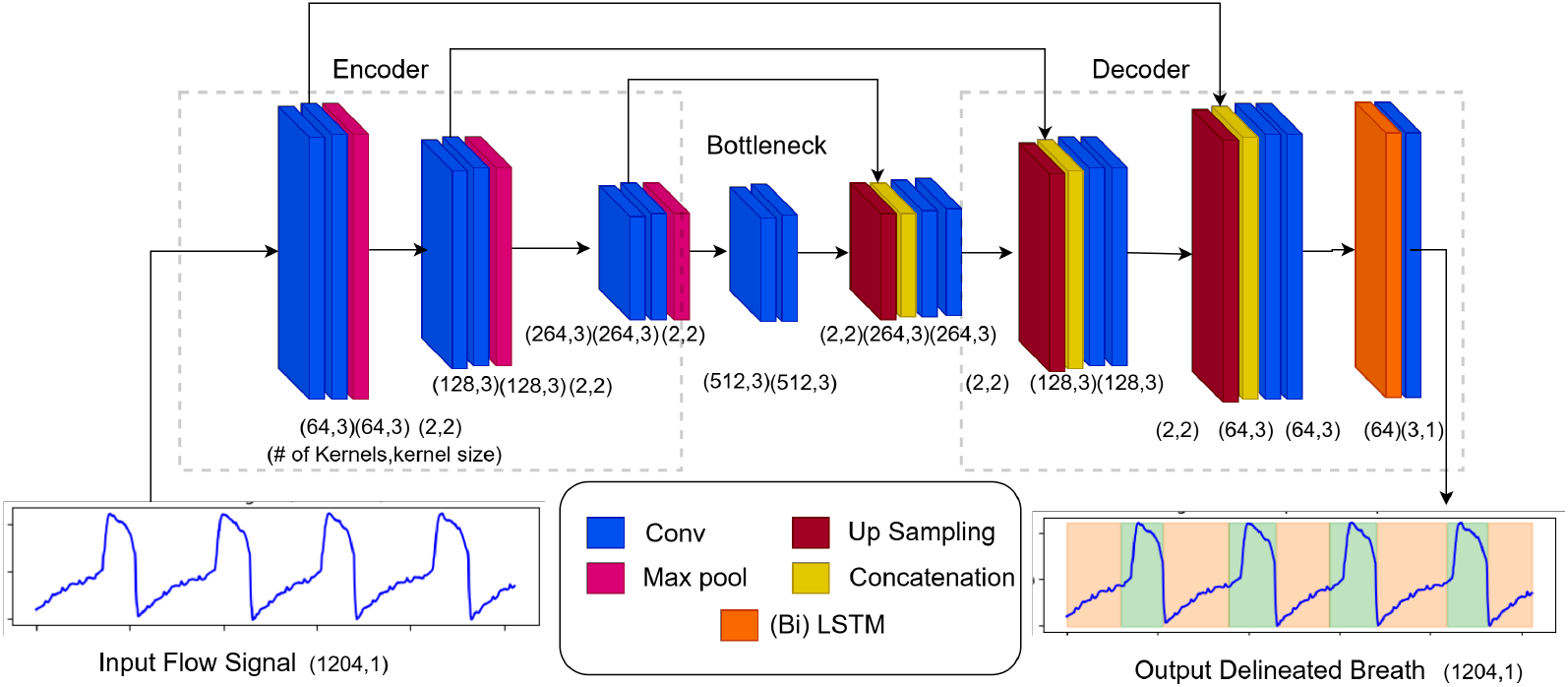
Architecture of the Convolutional Neural Network (CNN) - Long Short-Term Memory (LSTM) model for breath segmentation and delineation. This encoder-decoder structure utilizes convolutional layers for feature extraction, coupled with LSTM/Bidirectional LSTM (Bi-LSTM) layers in the decoder to capture temporal dependencies effectively.

### 3.5 Data Selection and Preparation

The training dataset was constructed from waveforms of patient PI0001, processed using the rule-based segmentation and delineation algorithms. From 20 hours of available recordings, we selected a representative subset of 7.8 hours, comprising approximately 7,000 breaths. This curation balanced computational efficiency with physiological diversity, incorporating both normal breathing patterns and VD events, including double triggering, flow limitation, and delayed cycling. Visual inspection ensured the exclusion of artifact-dominated segments while preserving waveform variability essential for robust model generalization.

Preprocessing involved three key stages: (1) artifact removal to eliminate significant signal gaps and non-physiological perturbations, (2) detrending to remove slow baseline wander and enhance respiratory phase transitions, and (3) filtering to suppress high-frequency noise while preserving breath morphology. The processed signals were partitioned into overlapping segments of 1,024 samples (≈ 16.4 seconds) with an 800-sample stride, maintaining temporal context while enabling effective data augmentation. Each patch was paired with its corresponding label segment, normalized to a consistent range. These input-label pairs were then organized and stored for training. Corresponding label segments were extracted from rule-based annotations and normalized to ensure consistent value ranges. To evaluate cross-patient generalizability, an independent test set (hereafter referred to as “groundtruth”) was prepared from patient PI0002. A 0.57-hour segment containing 533 breaths was manually annotated to provide a clinically relevant benchmark for assessment. This dataset included diverse breath types, including both normal and VD patterns.

### 3.6 Ground Truth Annotation and Validation Process

Manual annotations were performed on the independent test set to establish precise ground truth timing for inspiratory and expiratory phases, following the physiological criteria defined in Section 3.2. A custom MATLAB-based annotation tool was developed to ensure consistent and accurate marking of onset and offset events, recording both sample indices and timestamps. For performance evaluation, two complementary metrics were employed. The temporal accuracy was assessed through onset and offset timing errors:

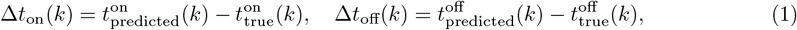

where 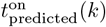 and 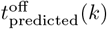 are the predicted onset and offset times for breath *k*, and 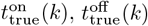 are the corresponding ground truth, respectively. The mean and standard deviation of these errors quantify systematic biases and random variations. Segmentation overlap measured via the Dice coefficient:

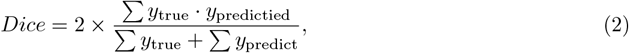

where *y*_true_ and *y*_predicted_ are binary vectors indicating ground truth and predicted phases. This metric effectively addresses variable segment durations and class imbalance inherent in respiratory phase analysis [45], unlike proximity-based metrics commonly used in ECG delineation [46, 47]. This dual-metric approach provides a comprehensive performance assessment where temporal errors quantify event detection precision, while the Dice score evaluates overall segmentation fidelity.

## 4 Results

### 4.1 Sensitivity to Parameters of Rule-Based Segmentation and Delineation

Figure 4 provides a qualitative assessment of the effectiveness of a rule-based algorithm for managing breath segmentation across different breath types.

**Figure 4.**
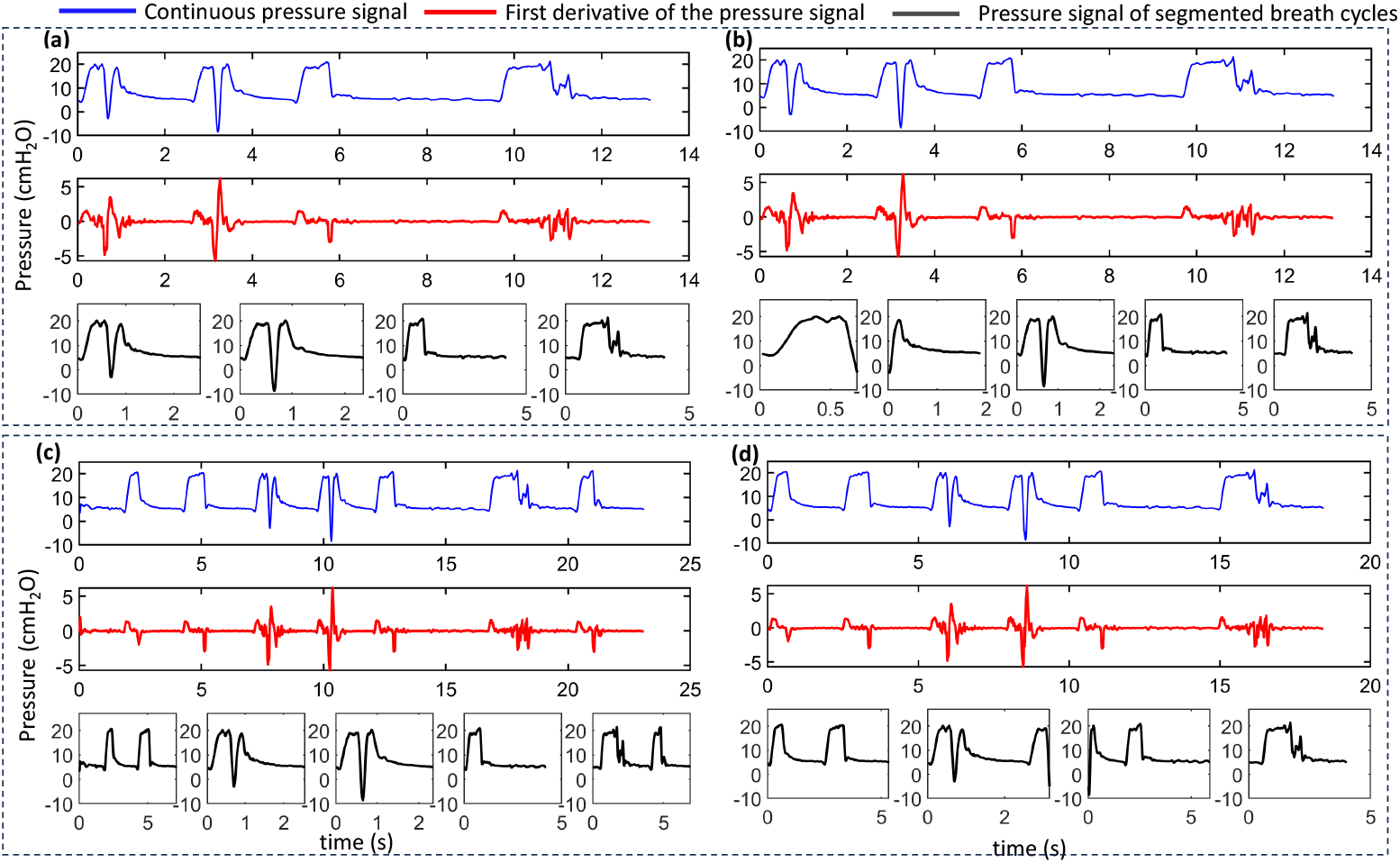
Effect of parameter tuning on rule-based breath segmentation. The first-derivative signal (red traces) is crucial for accurately identifying the start and end points of each breath (blue traces), thereby enabling the delineation of individual breath cycles (black traces). (a) Correctly detected breath boundaries with optimal parameters (*λ* = 0.7, *ξ* = 100). (b)-(d) Examples of incorrect segmentations resulting from suboptimal parameter values: (b) *λ* = 0.7, *ξ* = 40; (c) *λ* = 0.7, *ξ* = 160; and (d) *λ* = 1.4, *ξ* = 100.

Figure 4(a) shows breath segmentation using optimized parameters: a derivative threshold of *λ* = 0.7 and a minimum inter-breath interval of *ξ* = 100 samples (about 1.62 seconds). These values were determined through systematic validation to balance detection sensitivity and physiological relevance. The continuous pressure waveform (blue traces) illustrates the algorithm’s capability to identify breath boundaries, marked by black traces accurately. The parameter *ξ* plays an important role in managing the separation between consecutive breaths, helping to address irregular patterns and reduce false detections as shown in Figs. 4(b) and (c). If *ξ* is too low, the algorithm may mistakenly identify double-triggered breaths as separate events (Figure 4(b)). Conversely, increasing *ξ* to 160 samples can merge multiple breaths into one cycle (Figure 4(c)). The parameter *λ* influences detection sensitivity by filtering pressure changes based on their magnitude, with lower values enhancing the detection of subtle changes. In comparison, higher values may miss low-amplitude breaths during decreased effort or altered mechanics (Figure 4(d)).

Figure 5 demonstrates how parameter selection impacts breath phase delineation once breath segmentation is achieved.

**Figure 5.**
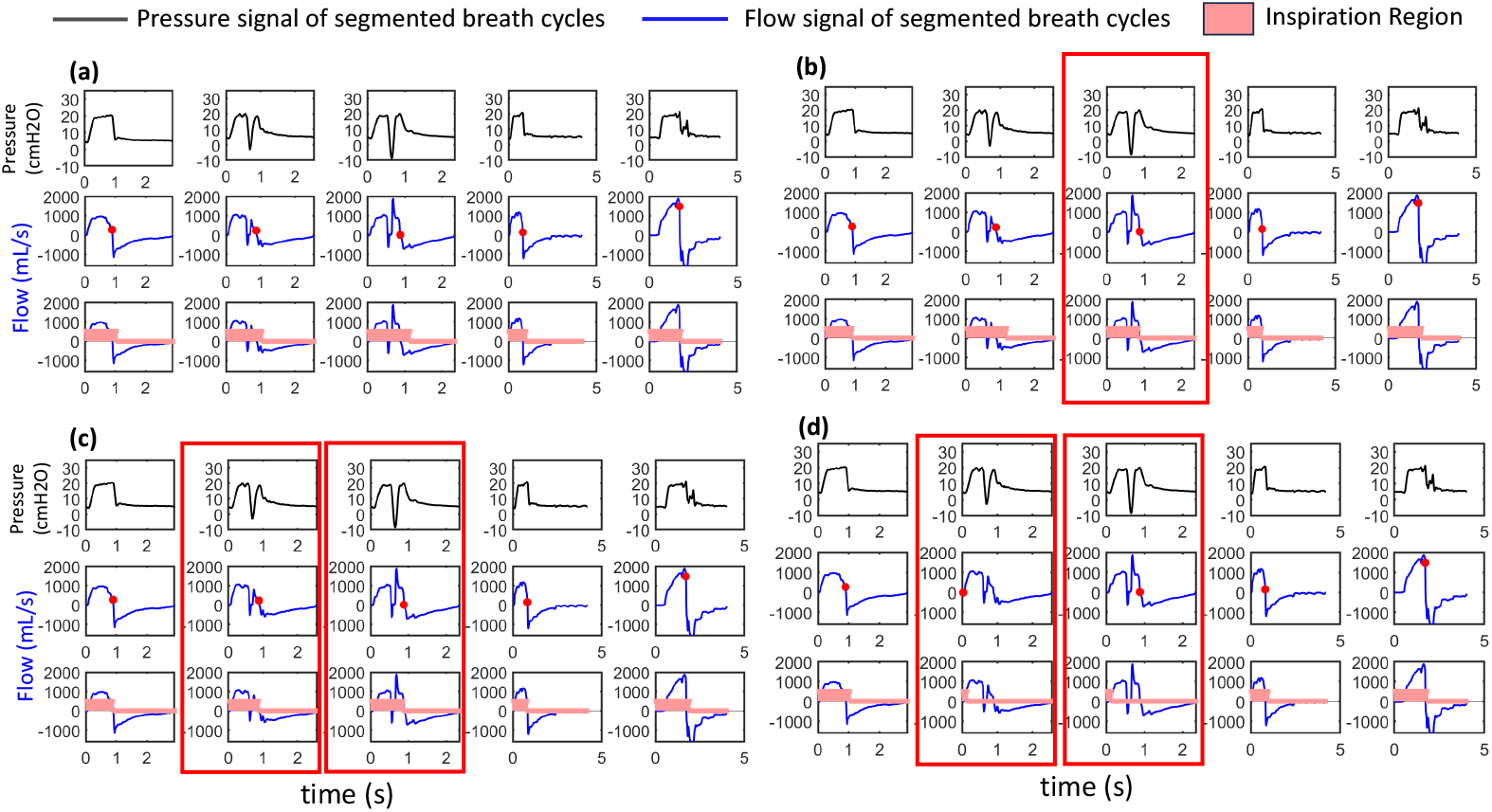
Each panel displays three aligned plots: the segmented pressure signal (black), the segmented flow signal with significant zero crossings (indicated by red dots), and the flow signal with the inspiration region shaded in red. The delineation results with various parameter settings are as follows: (a) *h* = 0.2, *w* = 5; (b) *h* = 0.8, *w* = 5; (c) *h* = 0.6, *w* = 8; and (d) *h* = 0.2, *w* = 15. Panel (a) displays accurate delineation, while panels (b) and (c) demonstrate issues with detection, highlighted using red rectangles.

In Fig. 5(a), the optimal parameters *h* = 0.2 and *w* = 5 enable precise identification of the beginning and end of the inspiration phase. In Fig. 5(b), keeping *w* constant while increasing *h* to 0.8 leads to misclassification of peaks exceeding this threshold as inspiration offsets, which is highlighted using a red rectangle. Although the significant zero crossings (indicated by red dots) remain unchanged, the higher value of *h* diminishes the detection of smaller peaks, resulting in missed inspiration offsets, particularly evident in the third breath of the subplot. Figure 5(c) illustrates that an increase in *w* to 8 does not affect the zero crossings, but setting *h* to 0.6 results in incorrect detection by filtering out smaller peaks. Meanwhile, Fig. 5(d) emphasizes the influence of adjusting *w* on zero-crossing selection. In this case, zero crossings within ±*w* samples are grouped, retaining only the first as significant. A larger *w* favors earlier crossings, which may not be physiologically relevant, thus impacting breath boundary detection. Therefore, selecting both peak height and window size parameters is critical for breath-phase delineation, as indicated by the moving red markers.

### 4.2 Quantitative Performance Evaluation of Rule-Based Method

Figure 6 evaluates algorithm performance against expert annotations across three dyssynchrony types: double triggering, delayed cycling, and early cycling [4], shown in Figure 6 (a). The inspiration and expiration phases are shown in green and beige, respectively. Figures 6(b) and (c) show successful phase delineation for delayed and early cycling events, indicating that the rule-based method can manage simple VD patterns. However, there are significant limitations with double-triggering scenarios. The algorithm fails to detect secondary inspiratory efforts, mistakenly merging them into expiration phases or misclassifying them as separate breaths (Highlighted in red boxes). These errors arise from fixed threshold limits that hinder adaptation to rapid flow-pressure changes typical of complex breathing. While the algorithm works well for simpler patterns, its rigidity with complex waveforms highlights the need for more adaptive methods in clinical respiratory monitoring.

**Figure 6.**
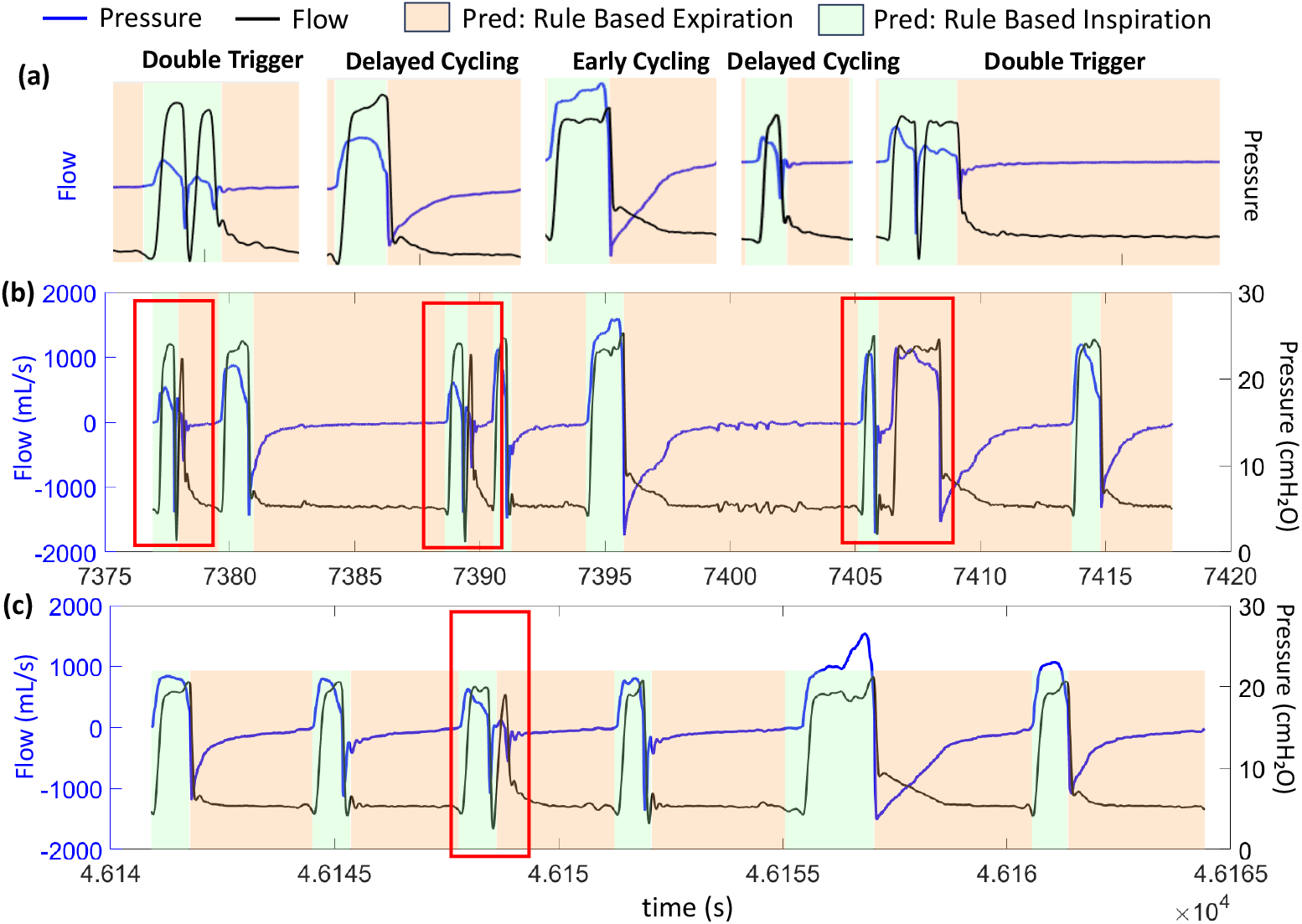
Comparative Analysis: Expert-Annotated Ground Truth vs. Rule-Based Algorithm Performance. Pressure and flow signals are depicted in blue and black, respectively, while the green and beige shaded areas indicate correctly identified inspiratory and expiratory phases. Panel (a) displays manually picked pressure and flow waveforms for three VD types: double-triggered, delayed-cycling, and early-cycling. Panels (b) and (c) display continuous respiratory waveform segments where the algorithm’s performance is scrutinized. Red boxes highlight specific VD instances where the rule-based approach fails to detect respiratory phase transitions accurately. Note that the x-axis shows relative time within extracted waveform windows, and not absolute time.

To quantitatively evaluate the performance of the rule-based method, we calculated the onset and offset errors along with the Dice score using the ground-truth test dataset (see Methods). The resulting Dice score of 0.9321 indicates strong correspondence with expert annotations. Table 1 presents detailed temporal error metrics, including an inspiration offset error of 0.007 ± 0.01805 s and an inspiration onset error of 0.01215 ± 0.03249 s. These metrics highlight the discrepancies between the outputs of the algorithm and the ground-truth test dataset. Variability in error values may stem from the characteristics of breath samples, with some breaths displaying notable misalignments. While the algorithm generally performs reliably, larger errors can arise in specific scenarios, such as during double-triggering events. Additionally, the method used to calculate the mean and standard deviation can influence the results, especially for smaller sample sizes, as errors are averaged across individual breath discrepancies. For example, with a sample interval of 0.016 s, most breaths may show small offset/onset errors (e.g., 40–60 samples corresponding to 0.64–0.96 s). In contrast, a limited number of breaths might exhibit larger deviations. Therefore, although the algorithm demonstrates consistency under normal conditions, reliance on static thresholds can limit accuracy in scenarios with variable dyssynchrony.

**Table 1:** Performance comparison of rule-based and deep learning approaches on the ground-truth test dataset. This table outlines the inspiration offset and onset errors, Dice score, number of tunable parameters, and training time for each method. The entries for inspiration offset and onset errors display the mean ± standard deviation.

| S. no. | Method | Inspiration Offset Error (s) | Inspiration Onset Error (s) | Dice score | # tunable parameters | Training time (s) |
| --- | --- | --- | --- | --- | --- | --- |
| 1 | Rule-based | $0.007 \pm 0.01805$ | $0.01215 \pm 0.03249$ | 0.9321 | $\lambda, \xi, w, h$ | - |
| 2 | Conv UNet | $0.004 \pm 0.015$ | $0.018 \pm 0.051$ | 0.9587 | 2,595,906 | 600 |
| 3 | Conv UNet + LSTM | $0.005 \pm 0.017$ | $0.017 \pm 0.050$ | 0.9588 | 2,628,930 | 640 |
| 4 | Conv UNet + Bi-LSTM | $0.004 \pm 0.013$ | $0.013 \pm 0.028$ | 0.9611 | 2,262,082 | 648 |

### 4.3 DL Model Training and Performance Benchmarking

The proposed U-Net architecture (Fig. 3) was trained using rule-based generated segmentation labels from Subject PI0001, which included 7,000 breaths segmented into temporal windows of (1024× 1 ×1) samples (approximately 16.38 seconds each). An overlap-based augmentation strategy was applied during training, using an 800-sample stride, while evaluation was performed on non-overlapping segments from the ground-truth test dataset. All models were optimized with the Adam optimizer and employed Dice loss to address class imbalance. Training durations ranged from 20 to 50 epochs, with a batch size of 16.

DL models show a clear advantage over rule-based methods for segmenting respiratory phases (Table 1). The Bi-LSTM-enhanced U-Net achieved the highest Dice score of 0.9611, which is a 3.1% improvement over the rule-based baseline. This improvement is due to the model’s ability to learn complex features instead of relying on fixed thresholds. The Bi-LSTM architecture was particularly effective, reducing inspiration offset errors by about 43% and onset errors by about 28%, while keeping error variability low (± 0.013 s for offset and ±0.028 s for onset). It accomplished this with greater efficiency, using 2.26 million parameters compared to 2.63 million in unidirectional LSTM models. Clinically, the Bi-LSTM model’s errors of less than 20 ms are well within acceptable limits, as ventilator trigger delays generally cause noticeable issues only when they exceed 100-200 ms. Even though training time increased slightly from 600 to 648 s, this is outweighed by the significant performance gains, making the architecture suitable for real-time processing. These results highlight the importance of bidirectional temporal context for accurately detecting respiratory events and suggest that the model captures essential physiological patterns rather than focusing on specific patient characteristics.

### 4.4 Visual Performance Analysis of Hybrid Bi-LSTM U-Net Model

Figure 7 presents a comparison of the performance between the U-Net and U-Net with Bi-LSTM models against the ground-truth dataset for normal breaths. The Bi-LSTM U-Net demonstrates improved sensitivity in detecting early inspiratory efforts (Segments S1–S3) and can identify subtle changes in airflow that are often overlooked by threshold-based algorithms. In Segments S4 and S5, the Bi-LSTM U-Net accurately recognizes the end of inspiration, whereas the baseline U-Net prematurely concludes the phase. Even in standard detection scenarios (Segment S6), the Bi-LSTM U-Net consistently outperforms the U-Net in identifying phase boundaries. This advantage is due to its bidirectional temporal processing, which enhances its understanding of respiratory patterns. Moreover, its ability to interpret early pressure changes in the correct temporal context allows it to overcome the limitations of fixed thresholds.

**Figure 7.**
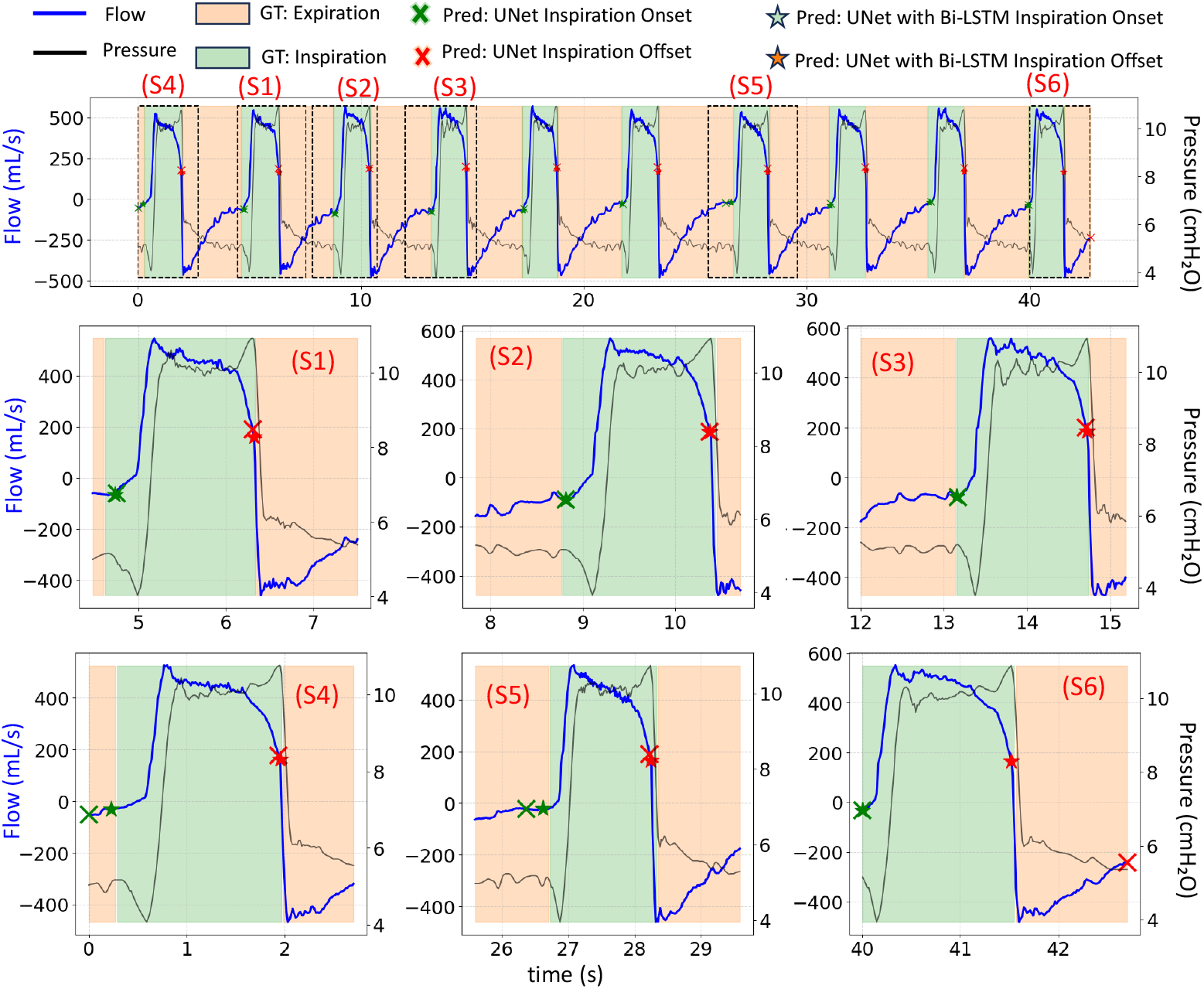
Comparison of ground truth and predicted breath phase delineation for the U-Net and U-Net with Bi-LSTM methods. The top panel displays a segment of respiratory data, highlighting flow (blue) and pressure (black). Green shaded areas represent ground truth (GT) inspiration, while orange areas indicate GT expiration. Predicted inspiration onsets and offsets are marked with a green “X” for the U-Net model and a white star outline for the U-Net with Bi-LSTM model. The six panels, labeled S1 to S6, offer zoomed-in views of individual breath cycles, illustrating both successful delineations and any discrepancies.

Figure 8 demonstrates the significantly improved capability of the Bi-LSTM U-Net model in detecting complex ventilatory patterns compared to traditional rule-based methods. The model accurately identifies inspiration onset, tracks stable breathing patterns, and adapts to transitional airflow changes, showing reliable performance in routine scenarios (Segments S1–S3). The model precisely detects the end of inspiration, a transition that rule-based methods often miss (Segment S4). In Segment S5, it correctly identifies a double-triggering event that the rule-based approach misclassifies, in which the VD breath is erroneously split into separate respiratory cycles. Similarly, in Segment S6, the Bi-LSTM U-Net model accurately captures the onset of inspiration, whereas a rule-based model misses it.

**Figure 8.**
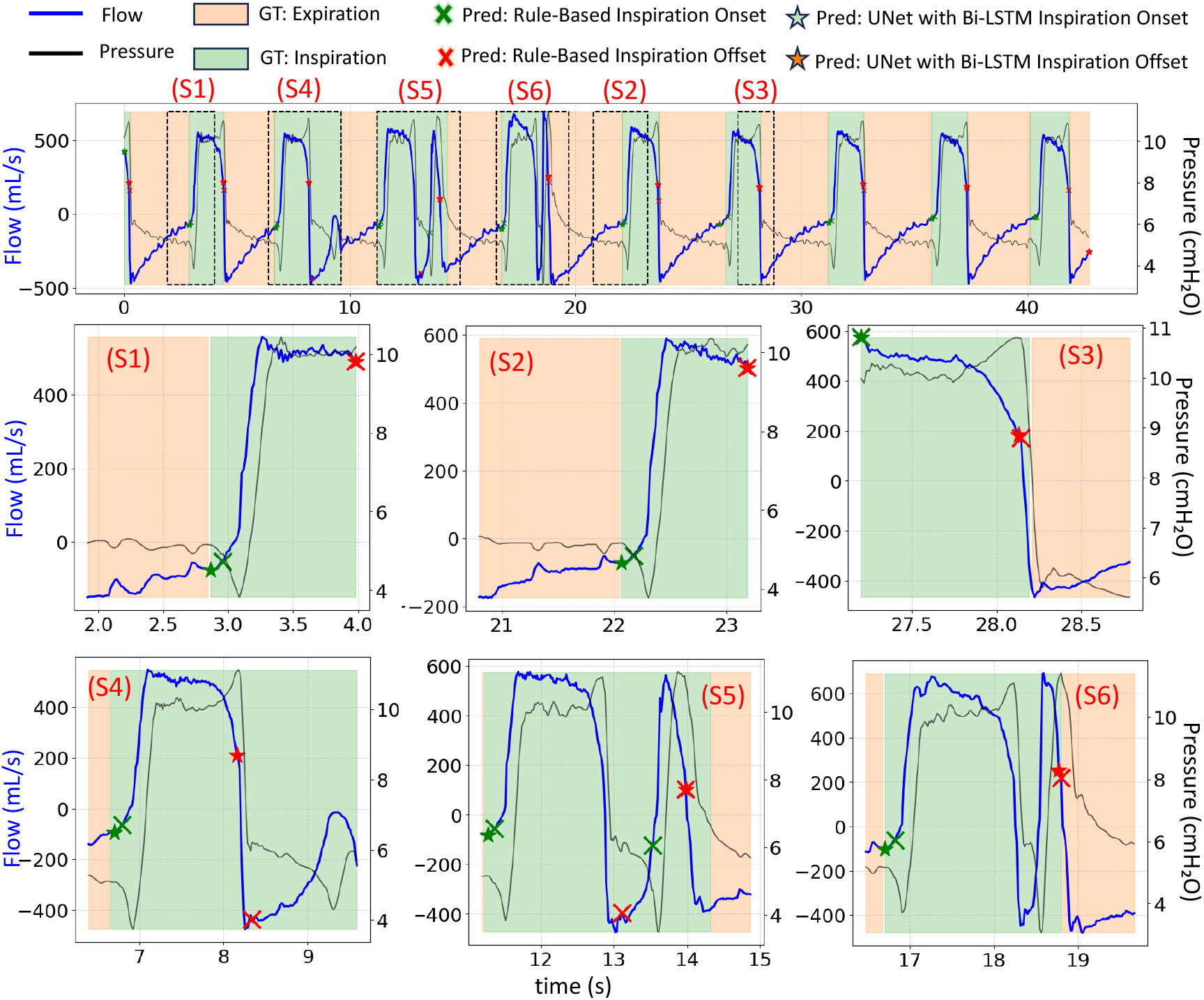
Comparison of ground truth and predicted breath phase delineation for the rule-based method and U-Net with Bi-LSTM methods. The top panel displays a segment of respiratory data, highlighting flow (blue) and pressure (black). Green shaded areas represent ground truth (GT) inspiration, while orange areas indicate GT expiration. Predicted inspiration onsets and offsets are marked with a green “X” for the rule-based method and a white star outline for the U-Net with Bi-LSTM model. The six panels, labeled S1 to S6, offer zoomed-in views of individual breath cycles, illustrating both successful delineations and any discrepancies.

## 5 Discussions

This study presents an automated pipeline for breath segmentation and phase delineation using a hybrid framework that integrates a rule-based method with a DL model. By combining adaptive, patient-specific rules to generate high-quality pseudo-labels for training, our approach overcomes the limitations of purely rule-based approaches, which struggle with waveform variability, and end-to-end DL models, which require extensive manual annotation. The Bi-LSTM-enhanced U-Net architecture demonstrates particular strength in capturing bidirectional temporal dependencies within respiratory cycles, enabling more accurate phase delineation than conventional methods.

Our results show that bidirectional recurrence consistently improves phase boundary precision, especially during VD events like double triggering, where conventional models underperform. The hybrid approach proves complementary: rule-derived labels provide scalable, physiologically plausible training targets, while the DL model corrects systematic rule failures and smooths label noise. The model achieved strong generalization to independently annotated data, indicating that learned features capture fundamental respiratory physiology rather than patient- or device-specific artifacts. In contrast, the rule-based method with fixed thresholds limits its ability to adapt to individual breathing patterns, potentially leading to missed transitions or inaccurate segmentation during complex respiratory phases.

Esophageal pressure (*P*_es_) is the gold standard for delineating respiratory phases, offering precise measurements of inspiratory effort [25]. It achieves near-100% accuracy in pinpointing the onset and termination of neural inspiration, reflecting the patient’s respiratory activity [25]. In contrast, non-invasive methods that infer phases from ventilator waveforms function on inference rather than direct measurement. This work proposes a method for phase delineation based on the characteristic signatures of pressure and flow signals to approximate *P*_es_-defined events. While effective for many breath types, its accuracy relies on clear waveform morphologies and is prone to failure in complex or pathological cases, such as severe air trapping, VD, or excessive noise, where physiological relationships may not hold. Future research should focus on developing a DL model trained on datasets containing both ventilator waveforms and synchronized *P*_es_ signals. This model would learn the complex mapping between airway pressure/flow patterns and *P*_es_-defined phase transitions. Once trained, it could serve as an inference tool for ventilator data lacking *P*_es_, potentially enhancing the accuracy and reliability of non-invasive breath segmentation and phase delineation, approaching the standards of invasive monitoring.

Our study has several limitations that warrant consideration. First, the relatively small dataset from a single institution and specific ventilator model (Maquet Servo-i) limits the generalizability of our findings. Future studies should validate these approaches across multiple centers and ventilator manufacturers. Second, while the Dice coefficient provides a valuable measure of segmentation overlap, it may not fully capture clinically relevant aspects of breath detection performance. The metric’s sensitivity to boundary definitions and potential insensitivity to certain types of temporal errors should be considered when interpreting clinical results. Third, the current approach requires careful parameter tuning for the rule-based component, which may not be feasible in all clinical settings. Future work should explore more adaptive thresholding techniques and potentially incorporate semi-supervised learning approaches to reduce dependency on rule-based labels. Fourth, we observed no improvement in onset error with DL models compared to the rule-based method. This may be because the inspiration onset can be influenced by factors such as patient variability, flow dynamics, and the subtlety of signal changes. If the DL models are not sufficiently trained on diverse and complex breathing patterns, they may struggle to detect these crucial onset transitions accurately. Finally, the computational complexity of the Bi-LSTM architecture suggests the need for further optimization. Future research should investigate model compression techniques, knowledge distillation, and more efficient architectural designs to enable more reliable breath delineation.

Despite these limitations, our findings demonstrate the significant potential of DL, particularly models with temporal modeling capabilities, to advance automated breath analysis in mechanical ventilation. The consistent performance across diverse respiratory patterns and the ability to generalize to unseen patient data suggest these methods could serve as valuable tools, reducing the manual effort required to develop reliable ventilator dyssynchrony detection algorithms.

## 6 Conclusion

This study introduces a fully automated hybrid framework for precise breath segmentation and phase delineation of ventilator waveforms. By synergistically combining a rule-based algorithm with a DL model, we address the critical limitations of manual annotation and the inflexibility of purely threshold-based methods. Our core contribution is the Bi-LSTM-enhanced U-Net architecture, which leverages bidirectional temporal modeling to achieve superior accuracy. The model attained a Dice coefficient of 0.9611 and demonstrated high temporal precision, with inspiration offset and onset errors of 0.004 ± 0.013 s and 0.013 ± 0.028 s, respectively. This performance represents a significant improvement over both the baseline rule-based method and standard U-Net architectures, particularly in handling complex dyssynchrony patterns like double-triggering, where conventional approaches fail. This hybrid pipeline provides a robust, scalable solution for automated patient-ventilator synchrony assessment, combining efficient rule-based pseudo-labeling with a DL model that adapts to waveform variability. Future work will focus on multi-center validation across diverse ventilator models and modes, computational optimization for potential real-time implementation, and the integration of this delineation framework into comprehensive systems for automated dyssynchrony detection and quantification.

## Data Availability

The data used in this study are not available

## Authorship Contribution

S.D.D. and D.K.A. conceived and designed the research, developed the mathematical model, and analyzed the data. A.D. and S.S. assisted with data curation. S.D.D. and D.K.A. evaluated the model and interpreted the results. S.D.D. prepared the figures and wrote the original draft of the manuscript. Both S.D.D. and D.K.A. edited and revised the manuscript, and they approved the final version.

## Acknowledgements

This work was supported through the Seed Grant (RD/0523-IRCCSH0-024) and the Institution of Eminence funding provided by the Indian Institute of Technology Bombay, India.

## Source code and dataset

The source code will be made available upon request.

